# Occupational Health Management in New Work – a Protocol for a Mixed-Method Study: Project BGM4NewWork

**DOI:** 10.1101/2023.09.21.23295923

**Authors:** Carla Rinne, Fiona Niebuhr, Anna-Sophia Wawera, Susanne Voelter-Mahlknecht

## Abstract

**Background:** The world of work is undergoing profound changes towards agile, flexible, democratic, and digital forms of work, so called *New Work* (NW). The COVID-19 pandemic accelerated these changes and confronted the working world with new challenges. Effects on employee health are ambivalent and remain unclear. Moreover, there is a lack of evidence as to whether existing occupational health management (OHM) measures meet the needs of employees working in new forms of work.

**Methods/Design:** This prospective mixed-method project will include four substudies to identify different NW forms, resulting health risk, benefits and protective factors in subgroups, and derive target group-specific OHM services. In the four substudies, the following methods will be used: (1) a scoping review, semi-standardized interviews, and an online survey, (2) a systematic review, an online survey, an expert workshop and qualitative interviews, (3) workplace observations, and (4) expert workshops. Recommendations for action will be derived from the findings of all substudies and summarized in a checklist for OHM in NW settings.

**Conclusion:** Findings will expand the state of knowledge about NW settings and associated health effects. The development of an evidence-based checklist for target group-specific identification of NW settings and associated health risks, benefits and protective factors can be used as a basis for action regarding OHM in companies. The findings can provide guidance on how future OHM services should be designed to meet the needs of employees.

## Introduction

Even before the COVID-19 pandemic, there have been transformations in the world of work towards agile, flexible, democratic, and digital forms of work, so called *New Work* (NW). NW is used as a term in multiple and heterogeneous ways; however, there is still no consensus on a uniform definition. It can be seen as a “container term”; a construct which is sometimes arbitrarily expanded or reduced [1]. Its original meaning can be traced back to Bergmann [2], who argued that people used to serve work. In the sense of today’s understanding of NW, however, work should serve the people. This “inversion” is central to understanding NW. Accordingly, NW can offer the opportunity to strengthen the resources of employees.

External, global, and local factors influence the NW transformation. In addition to globalization and climate change, the COVID-19 pandemic also acted as a catalyst for developments towards NW. Clear effects can be seen, for example, with regard to digitalization. Companies introduced NW measures, such as new digital and virtual tools, as this enables companies to improve their performance and actively adapt to the transformation process and external global factors [3]. A growing number of employees around the world were increasingly working in NW settings. This sudden change of work settings due to the pandemic was unprepared and led to various inconveniences for the workers such as unprepared desktop environment and a lack of information tools [4]. Furthermore, the COVID-19 pandemic led to increased locational flexibility, such as *working from home* (WFH). Prior to the pandemic, more than half of German employees had the option WFH—an opportunity that often remained unused [5]. The COVID-19 pandemic increased the proportion of employees WFH from 12% to up to 35% [6] and this trend continues—in 2021, 24.8% of the workforce in Germany worked from home occasionally and 10% daily [7]. The changes toward NW extend beyond the end of the COVID-19 pandemic. The trend for WFH apparently continues and will shape the future of work. In a study examining researchers’ experience of WFH, 66% want to increase their time WFH compared to their time WFH before the COVID-19 pandemic [8]. In summary, the pandemic has forced the world of work to make rapid changes towards NW, and this rapid change has led to longer-term movements towards more flexible and digital forms of work.

This movement in the world of work has a wide range of effects on employees and their health. Health effects resulting from new forms of work, such as flexible forms of work in terms of time and place, are the subject of controversial debates [9, 10]. In a systematic review, Lunde et al. [11] examined health outcomes of WFH including pain, well-being, overall-health, and burnout. They found both studies with positive effects and studies with negative effects on employee health. For example, Anderson et al. [12] examined, that employees reported higher positive affective well-being on days WFH. Furthermore, WFH was found to be associated with lower symptoms of depression in mothers with young children [13], as well as with a reduction in stress [14, 15]. Additionally, WFH can increase concentration and reduce work interruptions [16] and employees might experience an improved work-life balance while WFH [17]. On the other hand, Lunde et al. [11] also examined studies that found negative effects of WFH on health parameters. Song and Gao [18] reported an association of higher stress levels with WFH; other studies found no association between WFH and general health [19, 20]. In a study of German employees, a higher proportion of WFH was found to be associated with stress-related symptoms [21]. However, aspects such as high functionality of technical equipment while WFH showed positive effects on employee health [21]. Nonetheless, flexible work in the form of WFH can also increase back pain, lead to weight gain [22] and correlate with sleep quality [23] as well as psychosocial problems, e.g., cabin fever, isolation, and loss of concentration [24]. Moreover, other NW settings such as perceived flexibility at work can counterbalance negative effects of organizational, individual, and psychosocial factors of work-life balance [25]. Overall, there are ambivalent associations regarding the health of employees in NW settings; health-promoting as well as health-endangering effects are empirically supported.

*Occupational health management* (OHM) aims to maintain and improve the long-term health of employees at the individual and collective level [26]. OHM includes sustainable, systematic, and holistic measures of organizational and process design and takes all relevant interactions of work and health into account [27]. The spatial and temporal diversification of work (e.g., through WFH, coworking, and flexible working hours) as well as constant accessibility due to advanced digitalization pose challenges to the world of work and to OHM. Previous OHM approaches are not sufficiently tailored to forms of NW [28]. As the health effects of NW settings are not sufficiently studied, the requirements for the design of efficient OHM structures within these NW settings remain unclear. However, studies on these issues are essential, because most employees want to continue working in NW settings after the abrupt changes caused by COVID-19 and not return to pre-pandemic structures.

Examining health effects of NW and designing efficient OHM structures is a main aim of the project *BGM4NewWork*. OHM services should be demand-oriented. Because needs and risk factors are ambivalent across different NW settings and for different subgroups of employees, we will examine subgroups extracted from the empirical data using cluster analyses. The aim is to derive recommendations for action for OHM that are oriented toward the needs of employees in the corresponding subgroups and take into account the risks, benefits and protective factors of work in the specific subgroups of NW settings. The project BGM4NewWork is a third party funded consortium project with the partner UseTree GmbH (UseTree). UseTree focuses on the introduction of agile, human-centered work processes in companies and supports the development of holistic, innovative product solutions in the digital field. In addition, the project is supported by partners from cooperatives, health insurance companies, and business associations, among others.

The overall goal is to synthesize the data of all substudies of the project. This evidence synthesis will be used to develop a checklist based on subgroups of NW settings and associated health risks, benefits, and protective factors. This *BGM4NW-Checklist* will be used to derive suitable, needs-oriented OHM services. Accordingly, already implemented OHM services are compared with the current needs of employees in NW settings, and impulses for new or modified OHM services will be derived. We therefore aim to contribute to closing the described gaps in health care in NW settings. In addition, requirements and obstacles of OHM services and challenges of access and interfaces between and within sectors and providers will be identified. Based on an explorative mixed-method study design and thus on empirical foundation of qualitative (i.e., interviews and workshops) as well as quantitative data (questionnaire data), the following main research questions (RQ) will be addressed:

### RQ1a

Which subgroups of employees can be identified within NW settings? RQ1b: Which subgroups of NW settings can be determined?

### RQ2

What are the health needs and demands of different subgroups of employees and NW settings, and to what extent are there differences between these subgroups?

### RQ3

How should OHM access and services be designed in NW work settings?

### RQ4

How can OHM services and interfaces between and within sectors and OHM providers be improved in terms of insights into the health needs and demands of different subgroups of employees and NW settings?

## Method

We will conduct a prospective mixed-method design for our project, which is divided into four substudies (Fig 1) and is expected to take place between June 2022 and May 2025. Within the scope of our project, we focus on different effects of NW settings. Hence, we will identify associated health risks, benefits and protective factors in employee subgroups and finally derive recommendations for target group-specific OHM measures. All information provided here is based on the project plan. At the time of the publication of this study protocol, the project team was working on substudy 1.

**Figure 1.**
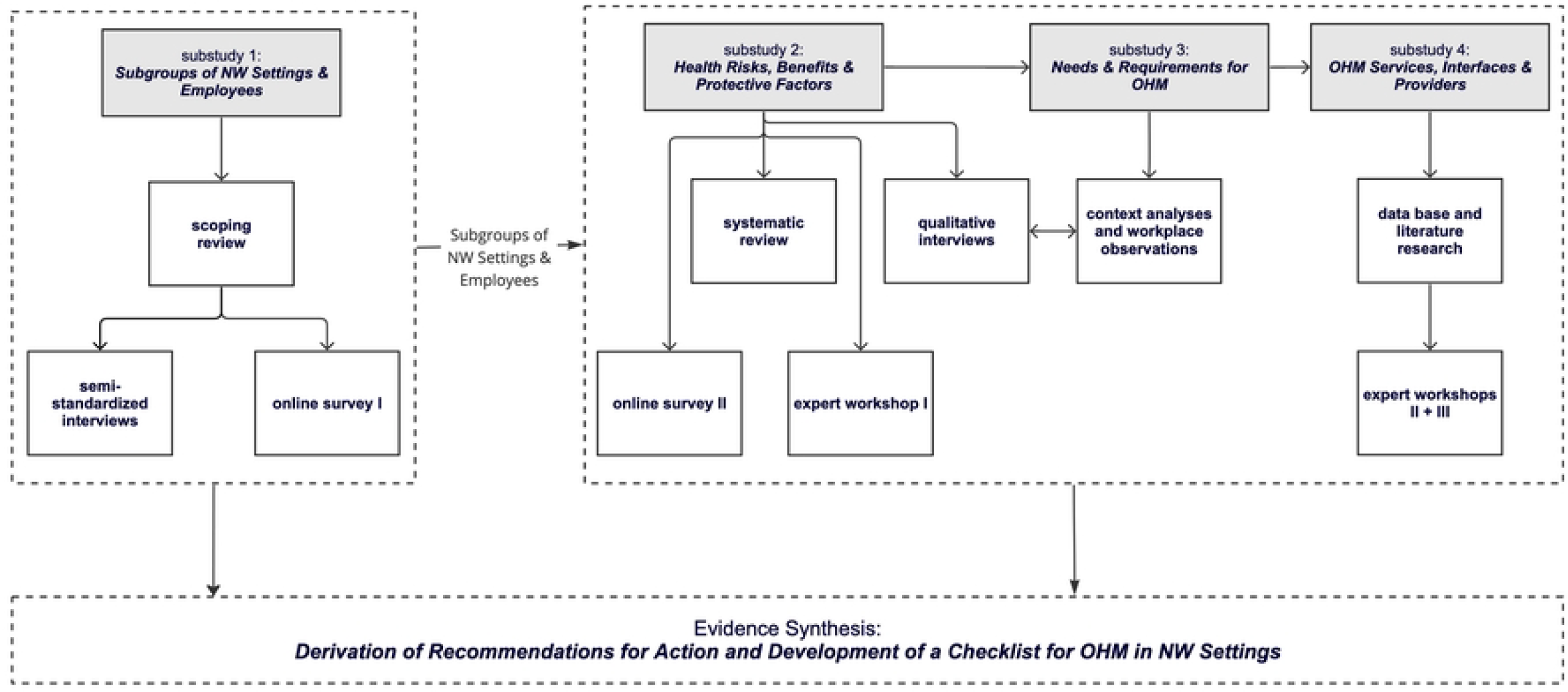
Overview of Substudies and Aim.

## Materials and methods

In the following, the study design, procedures, and recruitment of participants in the substudies will be described. The evidence of all substudies will be synthesized in order to achieve the overall goal of developing the BGM4NW-Checklist. Each substudy involves different methods and data collection methods. Furthermore, the aims of substudies and interdependencies will be examined.

### Substudy 1: Subgroups of NW Settings & Employees

The aim of substudy 1 is to address the first research question by identifying subgroups of NW settings and employees. Therefore, we postulate the following hypotheses:

#### H1a

Subgroups of NW settings can be identified.

#### H1b

Employee subgroups within NW settings can be identified.

To classify different subgroups of NW settings and employees, a *scoping review*, a *first online survey*, and *semi-standardized interviews* will be conducted.

The aim of the *scoping review* is to map out the most common settings of NW and the different subgroups of employees working in NW settings. The scoping review will follow the methodological framework of Arksey and O’Malley [29], which is one of the most commonly used frameworks for conducting scoping reviews in health and social sciences and will include studies from the databases Web of Science, Medline, PsycArticle, PsycInfo, Psyndex, and PubMed. All search strings will contain the following keywords “new work” or “work 4.0” and will be extended by AND connections; further search terms in German or English are “agility”, “flexibility”, “digitalization”, “empowerment”, “participation”, “digital leadership”, and “dissolution of boundaries”. Suitable synonyms of the search terms will also be researched based on piloting the search strings. All abstracts will be screened by the research team and following the PRISMA guidelines [30], relevant articles will be included. Data will be extracted for the type of study, author(s), title/year of publication, aims and objectives, setting and population group, and methodological design, and different forms of NW as well as different subgroups of employees working in NW will be mapped out.

An interview guideline for the *semi-standardized interviews* (*N* = 24) will be developed based on the literature extracted within the scope of the scoping review. Employees (*n* = 12) and decision-makers (*n* = 12; e.g., managers, work council) from companies with various forms of implemented NW work structures will be recruited for the semi-standardized interviews through the network of project partners.

Using the results of the scoping review, the *first online survey* will be compiled using various validated and established questionnaires and supplemented with own questions. The focus will be on the experience of new forms of work, the respective implementation in companies as well as opportunities and challenges within NW. The online survey will include questions about demography, work context, work activity, work culture, leadership, health, and new work. An a priori power analysis (G*Power Version 3.1.9.6) was conducted for an ANOVA to compare subgroups, resulting in a minimum sample size of *N* = 375. Our calculation included a small to medium effect (η^2^ = .04, *f* = .204), α = .05 and power = .9. To recruit participants for the online survey a panel company will be contracted. Working with a panel company ensures heterogeneous samples as well as guaranteed participation of the same employees at both measuring points. Since the questionnaire is a two-part questionnaire, the two measuring points are only two weeks apart and individuals with significant work-related changes, e.g., employees who changed their jobs, will be excluded. To identify subgroups of NW settings and employees within the quantitative data, a cluster analysis will be conducted. The cluster analysis will follow the methodological framework of Clatworthy et al. [31]. Participants working in companies with NW structures will be recruited. To consider all target groups, participants aged 18 to 65 years will be eligible to complete the survey. In particular, we aim to represent the group of over 55-year-olds in the data in order to be able to depict the age structure of the workforce in Germany. In addition, we assume that the group of over 55-year-olds has specific health risks and needs in the context of NW.

### Substudy 2: Health Risks, Benefits & Protective Factors

The aim of substudy 2 is to identify health risks and benefits of NW settings in order to examine RQ2. Therefore, we postulate the following hypotheses:

#### H2a

Subgroups of NW settings are associated with specific health risks, benefits, and protective factors.

#### H2b

Employee subgroups in NW settings have differentiated health risks, benefits, and protective factors.

Following the multi-method approach, a *systematic review* will be performed. The aim is to examine the relationships between a specific NW setting and specific health outcomes.

Furthermore, in substudy 2, the *second online survey* will be conducted. Participants from substudy 1 will be invited to complete the survey again via the panel company. The second online survey will include questions about demography, health, individual and organizational health associated risks, benefits, and protective factors. Some identical items will be used in order to control for changes between the first and second online questionnaire. Relevant health aspects and questions related to OHM will be added to the online questionnaire for the second survey.

In addition, substudy 2 will include *expert workshop I* with different stakeholders of OHM (*N* = 10), e.g., from occupational medicine, statutory pension, accident, and health insurance companies. The aim is to identify relevant NW settings and to explore what challenges the experts know from practice. The expert workshop will be used to identify initial findings on relevant health risks, benefits, and protective factors for working in NW settings as well as challenges for OHM. Participants will be recruited with the support of the project partners. The implementation will be based on focus groups; a guideline for the workshops will be prepared in advance.

Finally, exploratory *initial qualitative interviews* with employees (*N* = 35) will be conducted in substudy 2. Participating employees will be recruited from different NW settings with the support of project partners. The focus of the interviews is on health risks, benefits, and protective factors of NW settings. The results of the qualitative interviews will be used to derive initial target group-related criteria and objectives to be used in substudy 3.

### Substudy 3: Needs & Requirements for OHM

Focusing on RQ3, we will explore in the third substudy, how OHM access and services should be designed in NW settings by exploring the following hypotheses:

#### H3a

Employees in NW settings have specific needs in terms of OHM.

#### H3b

Influences and obstacles can be identified that have an impact on the access to and application of OHM.

The aim is to identify weak points between the needs of employees and the company’s offerings by comparing existing OHM measures with the current needs of employees, in order to derive impulses for new or modified OHM measures.

To identify the needs and requirements of OHM measures, *context analyses* and *workplace observations* will be conducted and use cases for practice will be derived by our consortium partner UseTree. Therefore, participants of the *qualitative interviews* (*N* = 35) of substudy 2 will be asked to participate in the workplace observations as well. The qualitative interviews and the workplace observations will be repeated after one year with the same participants in order to be able to map changes. We consider this important due to the velocity of the transformation processes in the current world of work.

### Substudy 4: OHM Services, Interfaces & Providers

Substudy 4 is designed to explore the fourth research question - how can OHM services and interfaces between and within sectors and OHM providers be improved? Hence, we postulate the following hypotheses:

#### H4a

Existing OHM services do not meet the specific requirements and needs of employees in NW settings.

#### H4b

Relevant OHM providers are not sufficiently integrated in terms of interfaces and offerings and networked across sectors.

Therefore, current OHM offers will be analyzed and discussed in *expert workshops II & III* in substudy 4. Interfaces and obstacles will be identified in a participatory approach together with the stakeholders. Participants of the expert workshop will be recruited via project partners. In expert workshop II (*n* = 10) company doctors, representatives of statutory pension, accident- and health insurance funds as well as experts in the field of human resources management from the involved project partners will participate. In expert workshop III (*n* = 10), health and safety representatives in NW settings, representatives of employer and employee associations and experts in the field of digitalized work from the project partners will participate.

Finally, all insights gained will be combined to develop the BGM4NW checklist. The results will be iteratively discussed with the project partners, processed and made available for their practice. The focus is on the promotion of health services in Germany.

## Data Analyses

All data will be processed by the research team. R-Studio and IBM SPSS Statistics (International Business Machines Corporation, Armonk, NY, USA) will be used to analyze the quantitative questionnaire data. For all analyses, the significance level will be set at 0.05. Outliers will be checked manually. Plausibility checks will be performed to ensure data quality. Descriptive and inferential statistical analyses will be performed to explore the research questions and test derived hypotheses. Cluster analyses will be used to synthesize the quantitative data to find subgroups of employees and NW settings. Qualitative data from interviews and workshops will be transcribed and subsequently analyzed using MAXQDA, following the approach of qualitative content analysis by Mayring [32]. The coding strategy combines deductive as well as inductive procedures.

## Monitoring and Ethics

The project has been submitted to and approved by the ethics committee of Charité – Universitaetsmedizin Berlin (No. EA2/189/22). In addition, a data management plan for the study (including all substudies) was developed and reviewed by the Clinical-Trial-Officers from Charité. Only the research team will have access to the original data. All participants generate a code.

Consequently, it is no longer possible to assign data to individuals after the data have been pseudonymized. All participants in all substudies will give their informed consent before data collection. The invitations to participate explicitly state that participation is voluntary and that non-participation or drop-out will not have any negative consequences.

The project will be conducted in accordance with the Declaration of Helsinki and the Guidelines of Good Epidemiologic Practice (GEP) of the International Epidemiological Association (IEA) and findings will be published in academic journals. Moreover, analyses and publications will be conducted in line with the European Code of Conduct for Research Integrity, Revised Edition (2017) and the Guidelines for ensuring good scientific practice.

## Conclusion

The project BGM4NewWork will follow a participatory multi-method design to develop an evidence-based checklist for target group-specific identification of NW settings and employee subgroups and their health risks, benefits, and protective factors. This aim will be addressed in four substudies, and the findings will be disseminated through publications in academic journals and multiplied by the involvement of project partners, i.e., OHM stakeholders. The knowledge of NW settings and employee subgroups, associated risks, accesses and barriers of OHM measures and the checklist will be used to maintain and improve the health of employees. Findings about target group-specific needs regarding OHM measures could inspire following intervention projects in the future.

## Data Availability

No datasets were generated or analysed during the current study. All relevant data from this study will be made available upon study completion.

## Acknowledgements

We thank Luara Severin dos Santos for her support in developing the concept of the project and Leonie Leitner for her support preparing the manuscript.

## Author Contributions

C.R., F.N., A.W. and S.V.-M. collaborated in the conception and implementation of this research project. C.R. and F.N. wrote the manuscript including figures. A.W. and S.V.-M. revised the manuscript. All authors reviewed, read, and approved the final manuscript.

## Availability of data and materials

To date, the data of the scoping review, semi-standardized inteviews, both online-questionnaires I and II are available. The further data collection has not yet started. Later, the data collected within this project can be requested from the corresponding author upon reasonable request. The data will not be publicly available, as this was assured to the participants in the study information as well as in the privacy statement.

## Ethics approval and consent to participate

The study protocol is in line with the current version of the Declaration of Helsinki (2013) and therefore approved by the Ethics Committee of the Charité – Universitaetsmedizin Berlin (No. EA2/189/22).

## Consent for publication

The article has not been published elsewhere. In addition, this manuscript is not considered for publication in another journal. All authors consent to the publication of this manuscript in PLOSone.

